# Clinically Interpretable Deep Learning via Sparse BagNets for Epiretinal Membrane and Related Pathology Detection

**DOI:** 10.1101/2025.06.05.25329045

**Authors:** Samuel Ofosu Mensah, Jonas Neubauer, Murat Seçkin Ayhan, Kerol Djoumessi, Lisa Koch, Mehmet Murat Uzel, Faik Gelisken, Philipp Berens

## Abstract

Epiretinal membrane (ERM) is a vitreoretinal interface disease that, if not properly addressed, can lead to vision impairment and negatively affect quality of life. For ERM detection and treatment planning, Optical Coherence Tomography (OCT) has become the primary imaging modality, offering non-invasive, high-resolution cross-sectional imaging of the retina. Deep learning models have also led to good ERM detection performance on OCT images. Nevertheless, most deep learning models cannot be easily understood by clinicians, which limits their acceptance in clinical practice. Post-hoc explanation methods have been utilised to support the uptake of models, albeit, with partial success. In this study, we trained a sparse BagNet model, an inherently interpretable deep learning model, to detect ERM in OCT images. It performed on par with a comparable black-box model and generalised well to external data. In a multitask setting, it also accurately predicted other changes related to the ERM pathophysiology. Through a user study with ophthalmologists, we showed that the visual explanations readily provided by the sparse BagNet model for its decisions are well-aligned with clinical expertise. We propose potential directions for clinical implementation of the sparse BagNet model to guide clinical decisions in practice.

## 1. Introduction

Epiretinal membrane (ERM) is a vitreoretinal interface disease characterised by the formation of a thin, fibrous layer of cells on the inner surface of the retina, particularly over the macula, the area crucial for central vision [8, 44, 34, 49]. Clinically, it manifests as a grayish, semi-translucent layer and can lead to various visual disturbances such as blurriness, distortion, and metamorphopsia (straight lines appear wavy) [25, 49]. The condition is most prevalent in individuals over the age of 50, with incidence reported within the range of 2.2% and 28.9% [17, 25, 49]. Commonly cited risk factors including advanced age, a history of ERM in the contralateral eye, and posterior vitreous detachment [1, 4, 34, 49]. ERMs may be asymptotic in the early stages, but can cause macular edema and other severe structural changes in the retina, as the disease progresses [3, 20, 25]. As the central part of the retina is normally affected, ERM can impact essential daily activities such as reading, driving, and facial recognition [14, 23]. As the disease progresses, ERMs can cause irreversible vision loss. Consequently, early detection – preferable in asymptomatic stages – is of great clinical importance [3, 23, 29, 51].

Three-dimensional (3D) Optical Coherence Tomography (OCT) is the current standard of care for detecting ERM [3, 21]. Such a volumetric scan essentially consists of multiple cross-sectional scans, known as B-scans. These can be used to create a 3D view of the macular region [16, 27]. Here, ERMs appear as a hyperreflective (bright) layer on the retinal surface. Even though OCT offers significant advantages in diagnosing ERM, the process of examining and interpreting the images is time-consuming for ophthalmologists [51, 31]. Deep learning models have been shown to accelerate ERM diagnosis from OCT scans, while also attaining high accuracy [11, 31, 36, 16, 24, 3, 54]. Despite these impressive results, a drawback of these models is their *black-box* nature, making them difficult for clinicians to interpret and trust [45, 53, 46, 22, 30, 52].

Recently, Ayhan et al. [3] curated the to date largest dataset representative of clinically relevant ERM cases and showed that DNNs can reliably detect ERMs of different severity in both the fovea and paracentral regions. To this end, the study used ensembles of DNNs in order to refine the decisions and post-hoc saliency maps obtained from individual models. The ensemble approach is computationally expensive and post-hoc saliency maps provide merely approximations of the underlying models’ decision-making processes [15, 33] and may be misleading [41]. The lack of faithful explanation of the inner-workings of deep learning models still poses challenges for their clinical adoption [30]. In contrast, inherently interpretable models that can readily provide insights into their decision making process are crucial for increasing trust, transparency and efficiency in clinical applications of deep learning [12, 13, 40, 42].

In this study, we used such a model called sparse BagNet [12], previously used to detect diabetic retinopathy from retinal fundus images, to detect ERMs from OCT images. This model combines the power of deep learning with an explicit representation of local evidence, reflecting the decision making process. Going beyond previous work, we also demonstrated the model’s capabilities in multitask settings, where we observed simultaneously good performance across various tasks involving determinants of ERM. Also, we evaluated the model’s ERM detection performance on external data, highlighting its generalisation potential for broader clinical application and performed a user study with ophthalmologists in order to establish the clinical relevance of sparse BagNet for ERM diagnosis.

## 2. Methods

### 2.1. Datasets

For model development and internal validation, we used OCT volumes from the Department of Ophthalmology at the University Hospital of Tübingen, which had been curated by Ayhan et al. [3]. The dataset consists of 624 OCT volumes that were acquired via Heidelberg Spectralis OCT from 461 patients. Overall, the dataset has 11^′^061 B-scans (on average 25 per volume) with a height of 496 pixels and varying widths. We removed all images with a width greater than 384 pixels (approx. 3% of the total) to ensure uniform size across the dataset.

All images were labelled by retina specialists for the presence of an ERM and its size (small (100-1^′^000µm) and large ERMs (*>*1^′^000µm)) and the quality of labels was also established through high inter-grader agreement (Cohen’s kappa scores of 0.963) [3]. For this study, we merged the small and large ERM classes, leading to 4^′^039 images without ERM and 6^′^683 images with ERM. A subset of images was further annotated for the following conditions commonly associated with ERM: Epiretinal fold (ERF) with 563 positive and 1^′^584 negative cases; foveoschisis (FOV) with 304 positive and 1^′^843 negative cases; intraretinal pseudocysts (IRP) with 1^′^019 positive and 1^′^128 negative cases; and epiretinal proliferation (ERP) with 534 positive and 1^′^613 negative cases.

To assess the generalisation performance of our model, we used a publicly available external dataset known as OCTDL [28]. OCTDL contains 2^′^064 images from 821 patients covering 6 retinal-related conditions, including ERM. We focus on 155 instances of ERM obtained from 71 patients. The images were captured using the Optovue Avanti RTVue XR camera [18], featuring 78 unique image dimensions with widths ranging from 616 – 1^′^158 and height ranging from 229 – 562.

We preprocessed the OCT images by first resizing them to 384 × 384, then normalised them by applying channel-wise mean substraction and standard deviation scaling. To prevent data leakage, we stratified the dataset by patient ID before splitting it into three sets: 75% for training, 10% for validation and 15% for testing.

### 2.2. Interpretable deep learning model for ERM detection

We used the sparse BagNet model [12] to develop an inherently interpretable DNN for ERM detection from OCT B-scans. The model is particularly well-suited for this task as its receptive fields are restricted to small regions in inputs and local information is processed first in order to capture fine-grained details in the vicinity. Accumulating local information across the input, the model then generates sparse class evidence maps, highlighting specific areas (individual patches) in the input that linearly contribute to the model’s final predictions. This approach has resulted in both high accuracy and inherent interpretability provided by the model [12, 13].

We trained the model using the internal dataset described above: 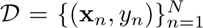, where each input image 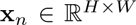 is a grayscale B-scan with height *H* and width *W*, and *y_n_* ∈ {0, 1}, with 0 and 1 representing the absence and presence of ERM, respectively. For a given pair of input image and its binary ERM label, the sparse BagNet model firstly processes small (33 × 33) overlapping patches in order to obtain an *M* × *M* × *D* feature map, where *M* is the height and width of the feature map, and *D* the number of feature maps. Then, it linearly aggregates local information from the feature maps towards a global prediction as per the original BagNet design [7].

However, the feature maps originally contain both positive and negative activations, suggesting that the model’s prediction is less influenced by input regions without clinical relevance.

The sparse BagNet addresses this by introducing an extra layer for constructing class evidence maps, which helps reduce the influence of clinically irrelevant regions. For *K* classes, it is of dimension *M* × *M* × *K*, which is obtained by processing the feature maps via 1 × 1 convolution. In addition, it is regularised by an *ℓ*_1_ term added to the cross-entropy loss (L_CE_) to penalise irrelevant activations. The regularisation promotes sparse visualisation and the total loss function is as follows:

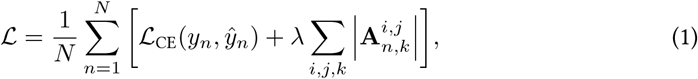

where 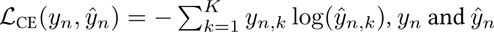 are respectively the true and predicted labels for **x***_n_*, *λ* denotes the sparsity parameter, and 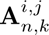 represents the activation at position (*i, j*) in the class evidence map for **x***_n_* and class *k*. Then, a spatial average pooling (SAP) is applied to the class evidence maps to generate logits. These logits are then processed through the softmax function to produce predictive probabilities of the model that estimates the presence of ERMs in the B-scans. The class evidence map can be upsampled and overlaid on the B-scans, offering inherently interpretable visualisations by highlighting clinically relevant regions in inputs (Figure 1). A significant advantage of the sparse BagNet model is its ability to provide such interpretability while delivering on par performance with black-box models [12, 13, 19].

**Figure 1:**
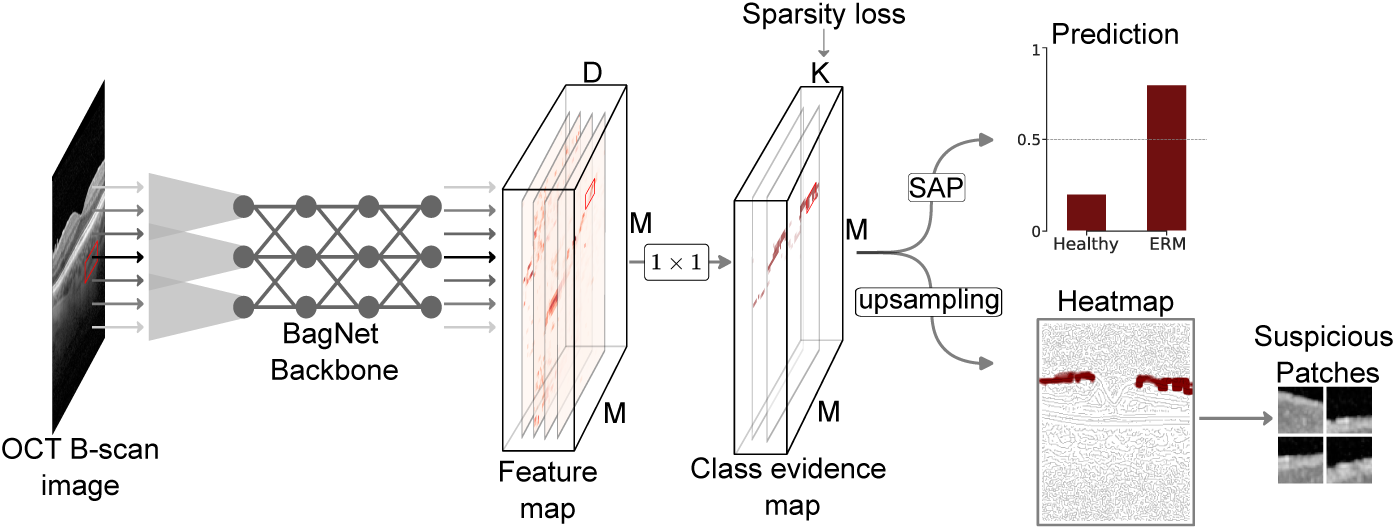
Overview of the sparse BagNet architecture for detecting ERM from OCT images and providing interpretations. The model first extracts feature maps from the input image, generates a class evidence map and employs a sparsity-constrained loss for learning. It provides logits by using spatial average pooling (SAP), which is fed to the softmax function for prediction probability. The class evidence map is then upsampled to create a heatmap to highlight regions of interest on the OCT image, along with extracting suspicious patches.

*Well-informed ERM detection via multitask learning.* ERMs are typically accompanied by ERF, FOV, IRP or ERP (see Section 2.1). In order to provide a more comprehensive support for the ERM diagnosis, we extended the learning scheme described above to a more complex scenario through multitask learning, for which the underlying model was modified to predict different outcomes simultaneously [9]. As a result of these changes, the new model shared hidden layers between all tasks, while maintaining different task-specific class evidence maps and prediction heads. This multitask model was then trained to jointly minimise all loss functions for the corresponding tasks. The loss function for the multitask is as follows:

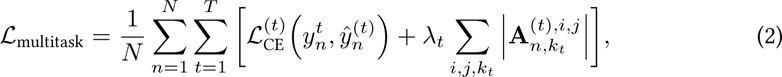

where *t* indexes the different tasks (from 1 to *T*), with each task having its own classification head. For each *x_n_* and task 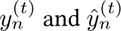 represent the true and predicted labels respectively, with 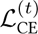 denoting the task-specific cross-entropy loss. The sparse representation for each task is promoted through task-specific regularisation parameters *λ_t_*. Each task generates its own class evidence maps 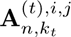, which represent the activation at position (*i, j*) for **x***_n_*, class *k_t_*, and task *t*. Thus, Equation (2) ensures that clinically irrelevant regions are minimised across all tasks while maintaining task-specific classification performance. Such a multitask learning approach was successfully used to predict the disease activity in the case of neovascular age-related macular degeneration (nAMD) along with its surrogate biomarkers, such as subretinal and intraretinal fluid, albeit with ensembles of more sophisticated DNNs that were not inherently interpretable [2]. To the best of our knowledge, we are the first to study multitask learning along with simple and built-in interpretability of BagNets, especially in the medical domain.

### 2.3. Experimental setup and implementation details

We used data random augmentation for model training to improve generalisation. We rotated the images within ±45 degrees, applied horizontal and vertical translations within ±30 pixels, adjusted brightness within ±10%, zoomed within ±10%, and flipped both horizontally and vertically. These augmentation techniques were applied to the images at random with a probability of 50%. In addition, we employed the *mixup* data augmentation technique [56] to enhance model robustness. *mixup* generates synthetic samples by linearly interpolating between pairs of samples (**x***_i_, y_i_*) and (**x***_j_, y_j_*) to create a new sample 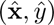 using the expression: 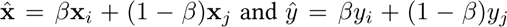, where *β* is drawn from a Beta(*α, α*) distribution. The *mixup* samples were drawn from the Beta distribution with *α* = 1.

We trained the models using the Adam optimiser [26] with a learning rate of 1 × 10^−4^ and weight decay of 5 × 10^−4^. With a batch size of 32, the models were all trained for 20 epochs. We used the same hyperparameter settings across all experiments to ensure comparability. In addition to sparse BagNet described in Section 2.2, we used ResNet–50 [19] and dense BagNet [7] for comparisons. The study was implemented using PyTorch (version 2.1.1) [37] as the primary deep learning framework, with additional libraries such as OpenCV [6] and sklearn [38]. We conducted computational experiments on the NVIDIA A40 GPU.

### 2.4. Design of user study

We conducted a user study at the University Hospital Tübingen with eight clinicians to assess the model’s ability to detect valid clinical evidence for ERM from OCT B-scans. From our internal dataset, we selected 40 images that the model predicted with high probability as containing ERM. Among these, 30 images were correctly predicted as containing ERM, while 10 were incorrectly predicted as containing ERM. It is important to note that the clinicians were unaware of which image was correctly predicted or not. Each image was presented to the clinicians with its corresponding heatmap overlaid on the OCT image. From each heatmap, at most nine distinct patches were extracted, representing regions of interest that the model considered most influential in its decision-making process (see Supplementary Figure S1). Each clinician independently assessed the potential evidence of ERM in the form of heatmaps and patches obtained from the sparse BagNet model.

For each patch, the clinicians were asked to evaluate and indicate whether they agreed with the model output. They could indicate “Yes” if they agreed that the patch contained evidence of ERM, “No” if they disagreed, and “Unsure” if they were uncertain. If “Unsure” is selected, a text box appeared, allowing clinicians to provide their reasons for their uncertainty.

We conducted the study using a web-based platform that was created using the Python web framework Django (version 4.2.1) and a secure PostgreSQL (version 15.3) backend database. The front-end of the platform, written in JavaScript, displayed an OCT image with its corresponding overlaid evidence. The numbered patches were placed below the OCT images with corresponding choice options. Users could also hover the cursor of the OCT image to see a magnified version. No personally identifiable information about the clinicians was ever shared; individual responses remained confidential; only aggregated results were reported in the study. The implementation of the platform is available at https://github.com/berenslab/retimgtools/releases/tag/v1.2.0.

### 2.5. Evaluation and statistical analysis

We assessed the model’s performance at detecting ERM using four metrics: accuracy, sensitivity, specificity and Area Under the Curve Receiver-Operating-Characteristics (AUC). We calculated confidence intervals by bootstrapping the data with 1^′^000 unstratified resamples. For the user study, we calculated a statistic that represents the agreement among clinicians. The statistic used is Randolph’s Kappa (*κ*), which is an extension of Cohen’s Kappa, designed to handle multiple clinicians responses [39]. It ranges from 0 to 1, where 0 indicates no agreement among clinicians and 1 indicates a perfect agreement.

## 3. Results

### Sparsity regularisation for ERM detection

First, we trained the sparse BagNet model to detect ERM from OCT images with varying sparsity regularisation (*λ*) and evaluated its performance on the test dataset (Figure 2). For an OCT image with ERM evidence, we found that a sparser model led to an evidence map highlighting smaller and localised regions, focusing specifically on clinically relevant retinal regions (Figure 2**A**). However, higher sparsity simultaneously reduced the model’s confidence, highlighting a trade-off between interpretability and model’s confidence. In contrast, lower sparsity produced higher confidence but with less localised heatmaps, which lacked precision. We observed that the intermediate sparsity setting (*λ* = 1 × 10^−5^) balanced this trade-off, preserving both good accuracy and meaningful interpretability. Similar effects were observed for OCT image without ERM evidence, reinforcing the interpretability-confidence trade-off (Figure 2**B**). Thus, *λ* = 1×10^−5^ was chosen for subsequent analyses of the study (Supplementary Figure S2; Figure S3).

**Figure 2:**
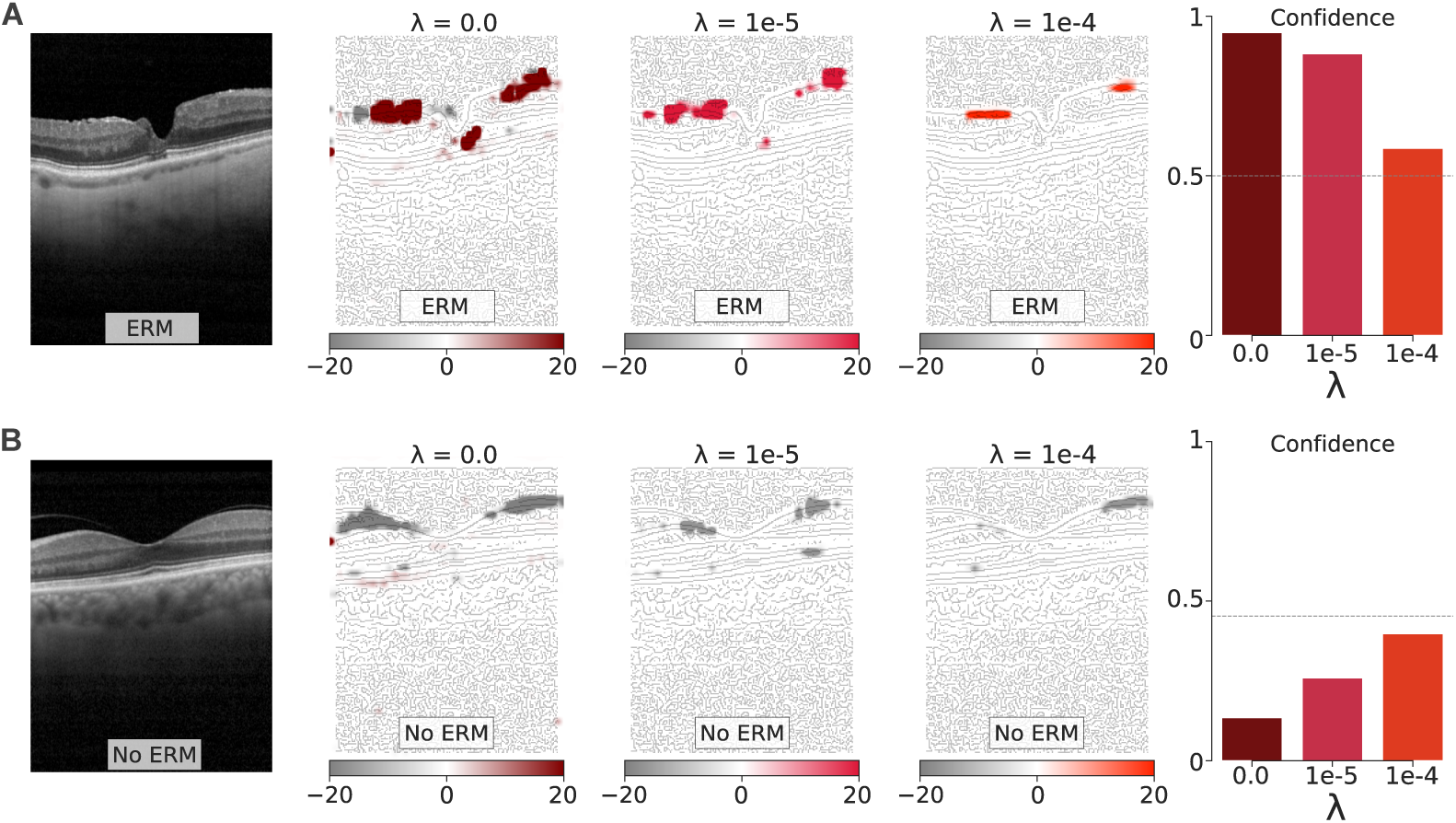
Visualisation of evidence maps of varying sparsity constraints with their corresponding confidence. Figure 2A shows evidence of disease in red, and they become less pronounced as *λ* increases. Figure 2B shows or almost no evidence of disease in OCT images. The confidence of ERM prediction also decreases as *λ* increases.

### Model comparison

Next, we compared the performance of the sparse BagNet model to the ResNet–50 and dense BagNet on the test set (Table 1). While the ResNet–50 achieved the highest accuracy at 94.46%, we found that the sparse and dense BagNet variants followed closely with accuracies 93.98% and 93.49% respectively. While the ResNet showed slightly higher sensitivity than the BagNet model, the BagNet variants had slightly higher specificity, likely influenced by the sparsity penalty that constrains activations. Thus, all model architectures showed comparable AUC, with the sparse BagNet providing the added benefit of inherent interpretability.

**Table 1:**
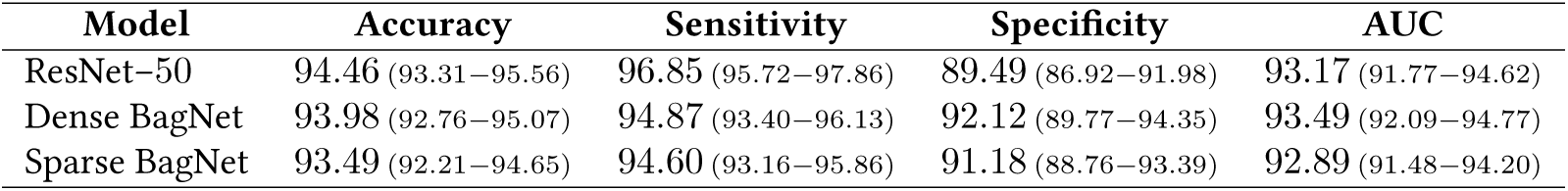
The performance metrics for Epiretinal Membrane classification on the test set.

### Attribution map comparisons

In addition, we evaluated attribution maps generated by the sparse BagNet, dense BagNet, and ResNet-50 models to visualise which regions in the OCT images were identified as disease-relevant (Figure 3**A**). The sparse BagNet model, which is inherently interpretable, produced highly localised class-evidence maps that targeted only the top layers of the retina – typically where the disease is present. In contrast, the dense BagNet model also highlighted relevant regions but extended its focus both above and below the retina, potentially capturing noise rather than purely disease-specific features. The ResNet–50 model, for which informative regions were identified via the post-hoc method GradCAM [43], generated coarser maps that highlighted larger regions of the OCT images, capturing various features but lacking the precision observed in the sparse BagNet maps. In addition, we provide examples of attribution maps generated by various post-hoc interpretation methods on the OCT images (Supplementary Figure S5).

**Figure 3:**
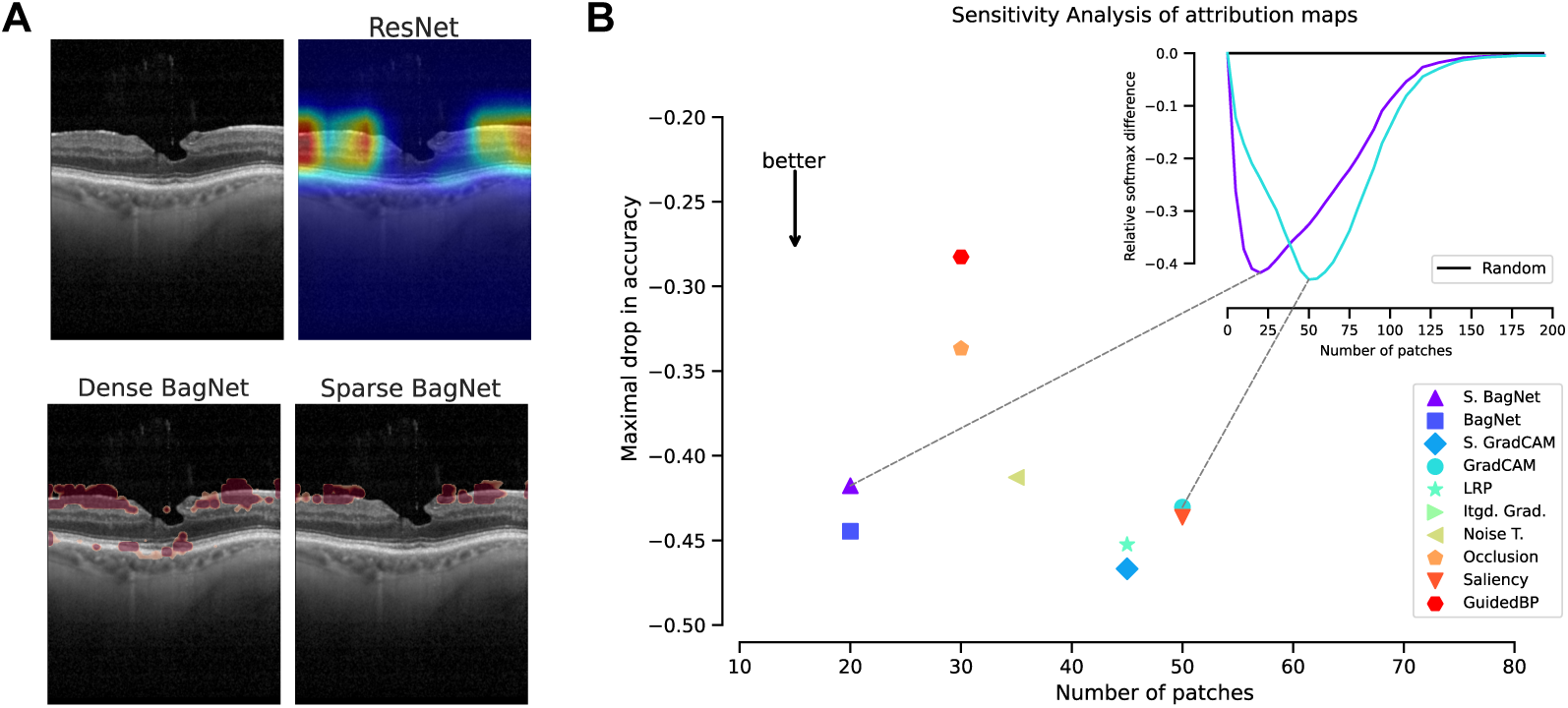
Comparison of attribution maps and sensitivity analysis. (**A**) Comparing the attribution maps of different models. The ResNet provides course evidence of disease, highlighting both the top layers and subsequent layers of the OCT image. The dense BagNet also highlights the top layers and subsequent layers but provides fine evidence of disease. In contrast, the sparse BagNet generates fine evidence of disease and only highlights the top layer of the OCT image. (**B**) A sensitivity analysis of multiple attribution maps, with points representing the maximum sensitivity change from a random baseline. Patches are progressively removed from the OCT images to highlight the effectiveness of identifying important regions in detecting ERM. The inset plot illustrates the complete sensitivity curves for two attribution methods along a random masking baseline (Supplementary Figure S4).

### Sensitivity analysis of attribution methods

To further assess interpretability, we conducted a sensitivity analysis to evaluate the localisations of regions of interest between the BagNet variants and other attribution map techniques (Figure 3**B**). The attribution maps used include Grad-CAM, smooth GradCAM (S. GradCAM) [47], layer-wise relevance propagation (LRP) [5], integrated gradients (Itgd. Grad.) [50], noise tunnel (Noise T.) [47], occlusion [55], saliency, and guided backpropagation (GuidedBP) [48]. In the analysis, we only included OCT images containing ERMs to specifically evaluate the ability to identify regions of interest that are critical for detecting ERMs by various attribution map techniques. We systematically perturbed the images by progressively replacing top-ranked patches of size 33 × 33 (extracted from the highlighted regions generated by the attribution map methods mentioned above) with zeros. In addition, we established a baseline by randomly replacing patches with zeros in the images. We then calculated the maximum change in performance between the baseline and the attribution map methods, allowing us to evaluate the effectiveness of each method in identifying regions of interest within the images. We found that perturbing images according to the heatmaps obtained from the sparse and dense BagNet achieved a large performance drop, requiring fewer patches than other models, highlighting their efficiency in identifying critical regions in the images (Figure 3**B**). In contrast, the other attribution map methods, required more patches to achieve comparable sensitivity changes, suggesting a more diffuse focus across the image.

### Generalisation to an additional external data set

The sparse BagNet model also demonstrated strong generalisation capabilities by achieving an accuracy of 89.03% in detecting ERMs on an additional external dataset OCTDL (see dataset description in Section 2.1). Notably, their heatmaps highlight disease-relevant areas, focusing specifically on the top of the retina, where ERMs are known to manifest (Figure 4; Supplementary Figure S6). This underscores the model’s potential clinical applicability across diverse datasets.

**Figure 4:**
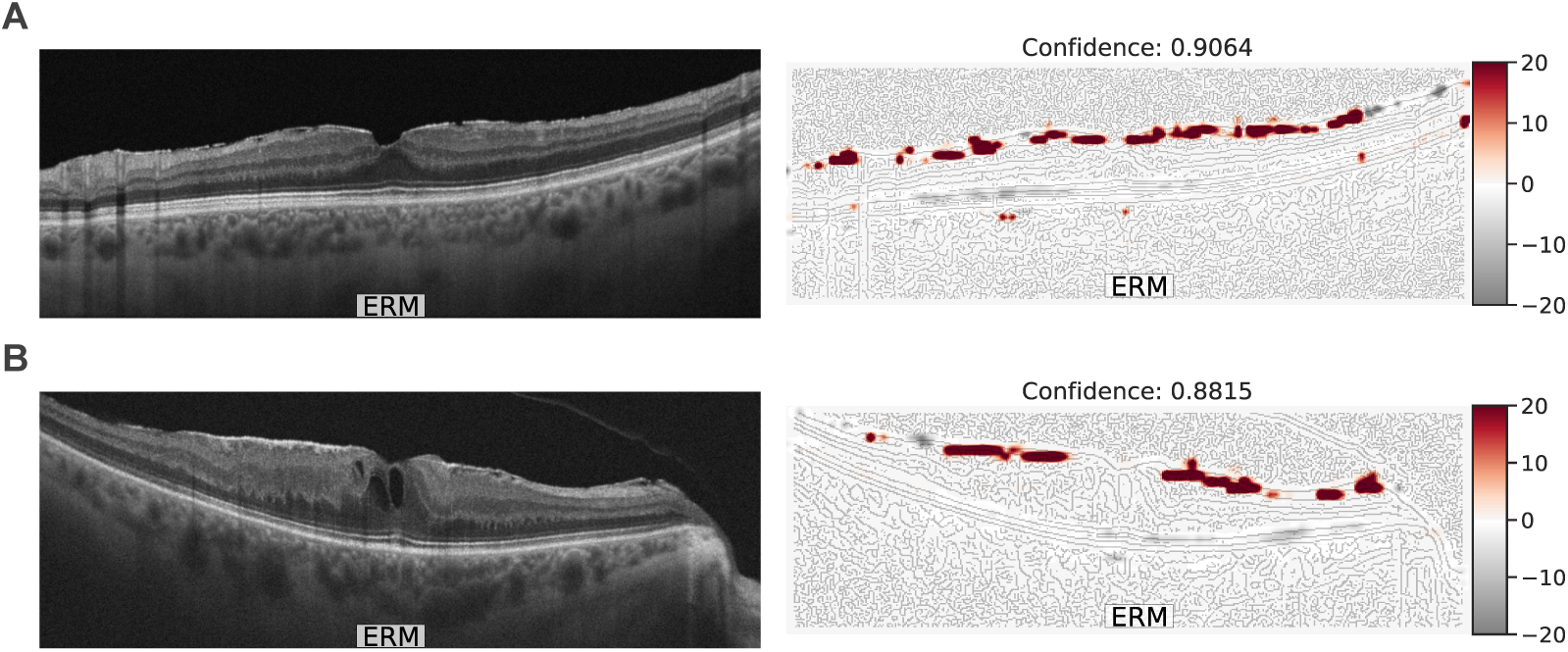
Evaluating the model on an external dataset. In the examples shown in this figure, the model correctly predicted the images with high confidence and highlighted suspected regions of interest.

### Multitask learning for detecting ERM-related pathologies

We extended sparse BagNet to multitask classification of ERM-related pathologies including ERF, FOV, IRP, and ERP (Table 2). This multitask capability is clinically significant because patients often present more than one co-occurring condition [3]. The multitask sparse BagNet model consistently demonstrated excellent performance across all evaluated ERM-related pathologies. The model achieved exceptionally high accuracy for foveoschisis (FOV) and closely followed by Epiretinal fold (ERF), with lowest accuracy for Epiretinal proliferation (ERP). The sensitivity values were all above 90% with the exception of FOV, indicating a high detection rate for ERM-related pathologies. The specificity was more variable between ERM-related pathologies, with FOV demonstrating the highest specificity and ERP the lowest. As a consequence, the model achieved excellent AUC scores, which provide a balanced measure of model performance, across all pathologies, with ERF achieving the highest AUC. Again, ERF showed a well-balanced performance with high scores across all metrics. Also for the multi-task model, we overlaid the class evidence map for individual pathologies on the OCT images with different colors corresponding to different pathologies (Figure 5). We found that the multi-task heatmaps often indicated mostly highlighted regions in which the disease should be found: For example, an intraretinal pseudocyst was identified within the tissue (Figure 5A), while a foveoschisis was found in a rupture of the fovea (Figure 5B). Furthermore, although ERM, ERF and ERP all occur near the retina’s surface, the results showed the model was able to distinguish these subtle differences [10, 32, 35]. The results obtained for the multitask sparse BagNet emphasise the model’s ability to learn in a multitask manner and do so with high confidence (Supplementary Figure S7).

**Figure 5:**
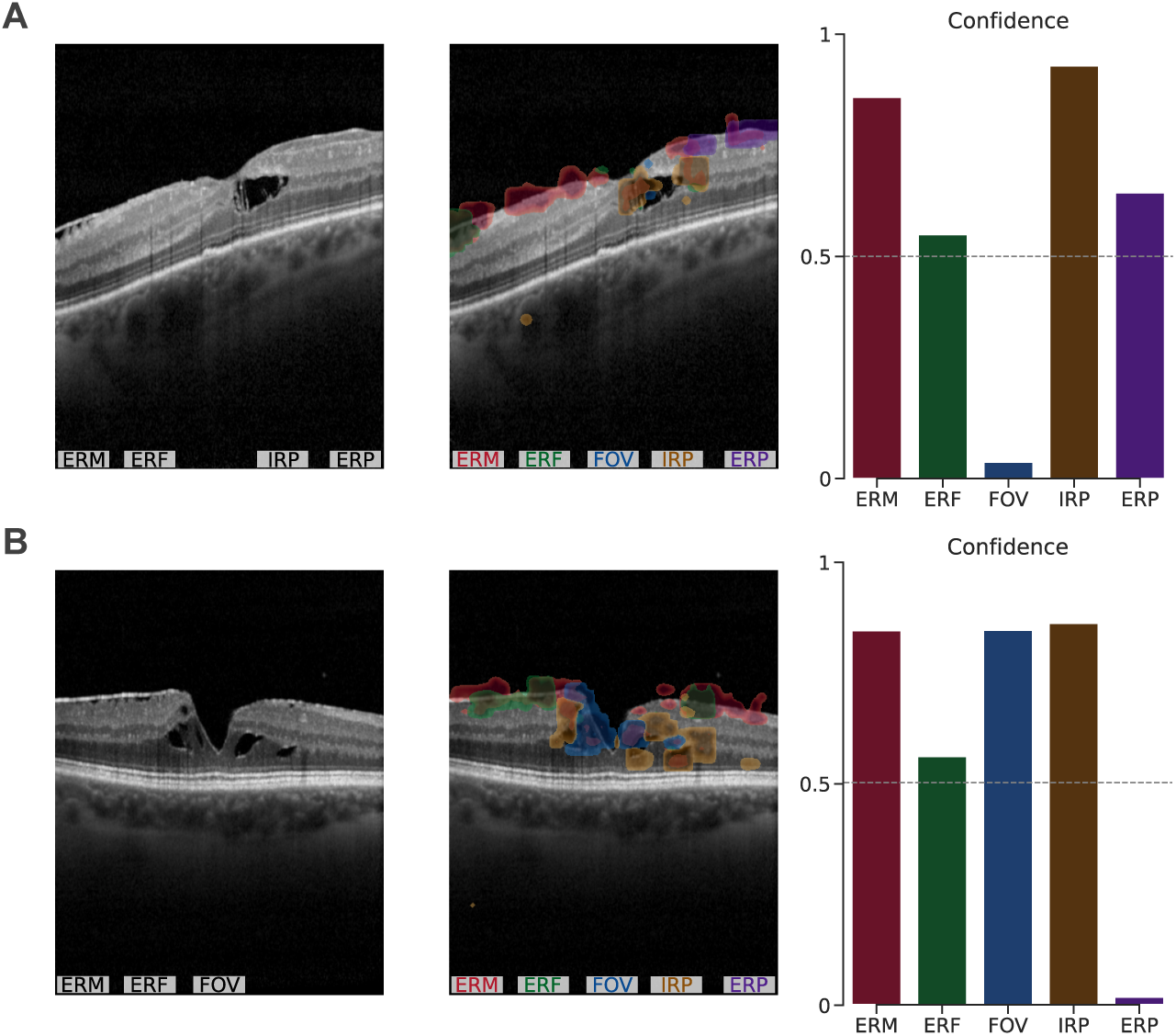
A multi-task evaluation of the sparse BagNet model. The model can accurately predict and provide evidence for each pathology with high confidence. The grey text in the first column represents the presence of pathology in the image, while the coloured text in the second column corresponds to the evidence generated by the model.

**Table 2:** Performance metrics of multitask sparse BagNet for different pathologies.

| Pathology | Accuracy | Sensitivity | Specificity | AUC |
| --- | --- | --- | --- | --- |
| ERM | 91.24 (89.89–92.59) | 92.19 (90.62–93.58) | 88.59 (85.75–91.69) | 90.39 (88.74–92.07) |
| ERF | 95.73 (93.45–97.72) | 96.23 (90.24–100.0) | 95.64 (93.22–97.89) | 95.93 (92.93–98.20) |
| FOV | 97.15 (95.44–98.86) | 85.71 (75.43–95.16) | 99.01 (97.73–100.0) | 92.36 (87.17–97.14) |
| IRP | 90.03 (86.61–93.16) | 93.05 (89.01–96.57) | 86.59 (81.20–91.62) | 89.82 (86.51–92.96) |
| ERP | 85.19 (81.48–88.60) | 92.73 (87.72–97.25) | 81.74 (76.67–86.56) | 87.24 (83.69–90.59) |

ERF - Epiretinal Fold, FOV - Foveoschisis, IRP - Intraretinal Pseudocyst, ERP - Epiretinal Proliferation

*User study evaluation.* Finally, we conducted a clinician-based evaluation of patches identified by the sparse BagNet model from OCT scans (Figure 6). We observed that for patches derived from true positive images, most were correctly identified as “Yes”, indicating that distinct features in these patches supported confident diagnosis. However, a subset still received “No” or “Unsure” labels, suggesting that subtle presentation of the disease could hinder clear identification. Notably, even among the false positive images, most patches were also labelled “Yes”, though a small fraction were deemed “No” or “Unsure”, reflecting occasional ambiguity. Overall, the distribution of responses underlined both the clinicians’ high sensitivity in recognising ERM where it was truly present and their recognition of potential disease features in images initially deemed false positives – implying that subtle pathological cues may have been overlooked in the original labelling (the 10 false positives are shown in Supplementary Figure S8).

**Figure 6:**
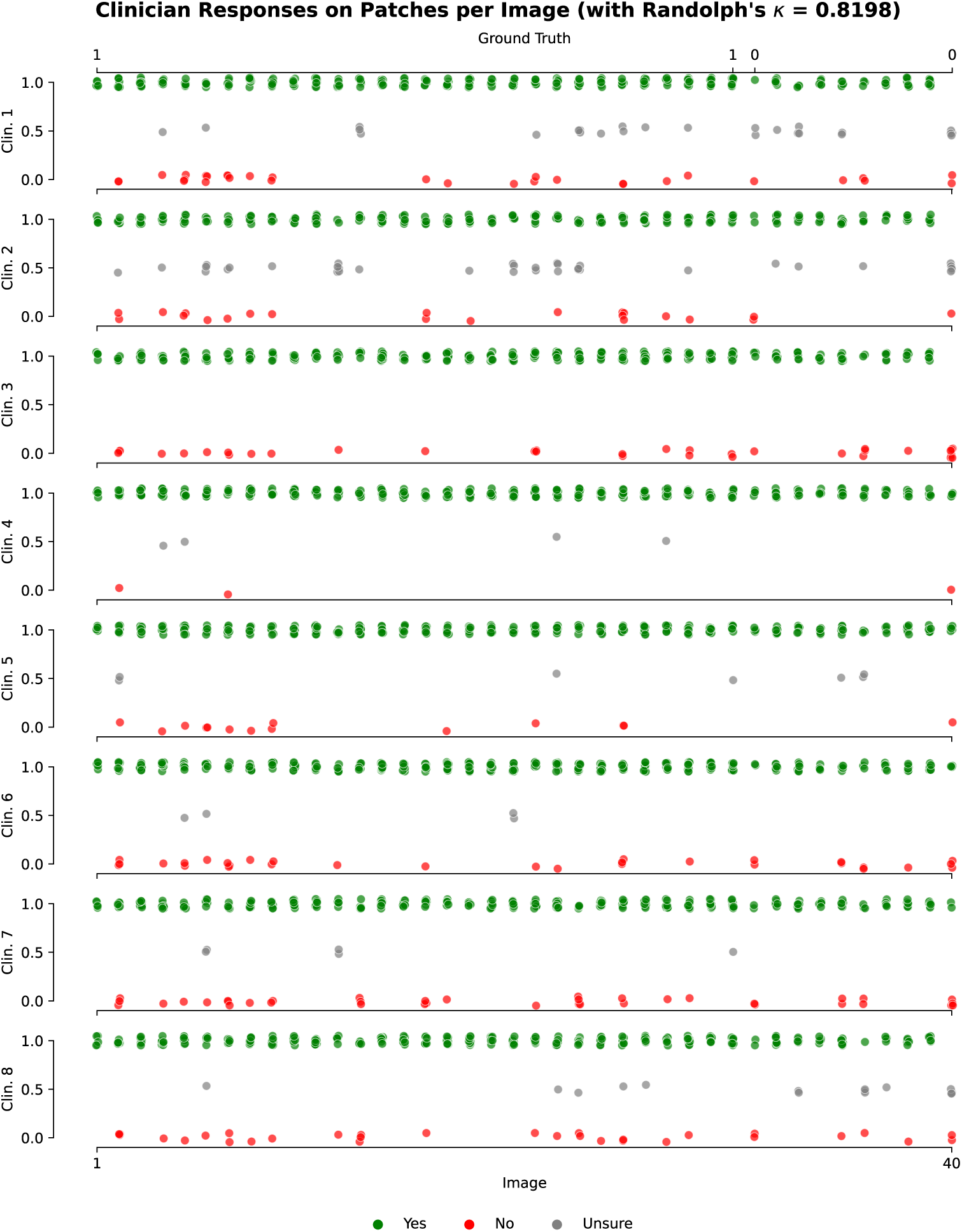
User study evaluation. Clinicians’ patch-level classifications of potential ERM in 40 OCT B-scan images (30 true positives, 10 false positives). Each dot represents a single clinician–patch assessment (Yes, No, or Unsure), illustrating a substantial inter-rater agreement.

Moreover, the clinicians provided free-text notes, reflecting the instances in which they selected “Unsure” on whether a patch contained ERM or not. Overall, there were 83 instances where participants entered free-text. Several participants remarked on the subjective nature of labelling borderline findings (e.g. “depends where you make the cut-off”, “cannot clearly be separated from inner retinal layers”), while others raised potential methodological or labelling issues. In general, the notes suggest that uncertainties in the user study most frequently arose from one of four issues: very small or subtle lesions (37 occurences), hyperreflective points challenges (24 occurences), difficulty separating the membrane from the retinal surface (13 occurences), and occasional concerns about image quality (5 occurences). Four additional comments were not specifically medical in nature, instead referencing unusual image configurations or missing display labels. Moveover, the inter-rater agreement was notably high with a Randolph’s *κ* = 0.8198, indicating a substantial agreement among the clinicians.

## 4. Discussion

This study presents a new inherently interpretable model for ERM detection including the reliable detection of ERM-related pathologies, despite only trained on image-level labels. Our results underline the dual benefits of the sparse BagNet, that is, attaining on par performance with state-of-the-art models and utilising sparse activations to highlight only regions of the retina that are clinically relevant. By focusing on the top-layer of the retina – where ERMs are known to manifest – the model enhances trust by producing fine-grained evidence maps that closely mirrors the decision-making process of ophthalmologists and can help to aid ERM detection and detection of ERM-related pathologies. Despite being trained on 2D images, in clinical practice, such a model could be used to process a patient’s OCT volumetric data slice by slice (comprising 25 B-scans). This would result in volume annotations, which can be shown in 3D to visualize regions covered by ERM or regions in which other pathologies are present (Figure 7) to aid clinical decision making. A major advantage of the model is the control over sparsity facilitated by the regularisation parameter (*λ*), which allows it to balance predictive power with interpretability. This inherent interpretability provides a significant advantage over conventional CNN-based models, which often require post-hoc interpretable methods.

**Figure 7:**
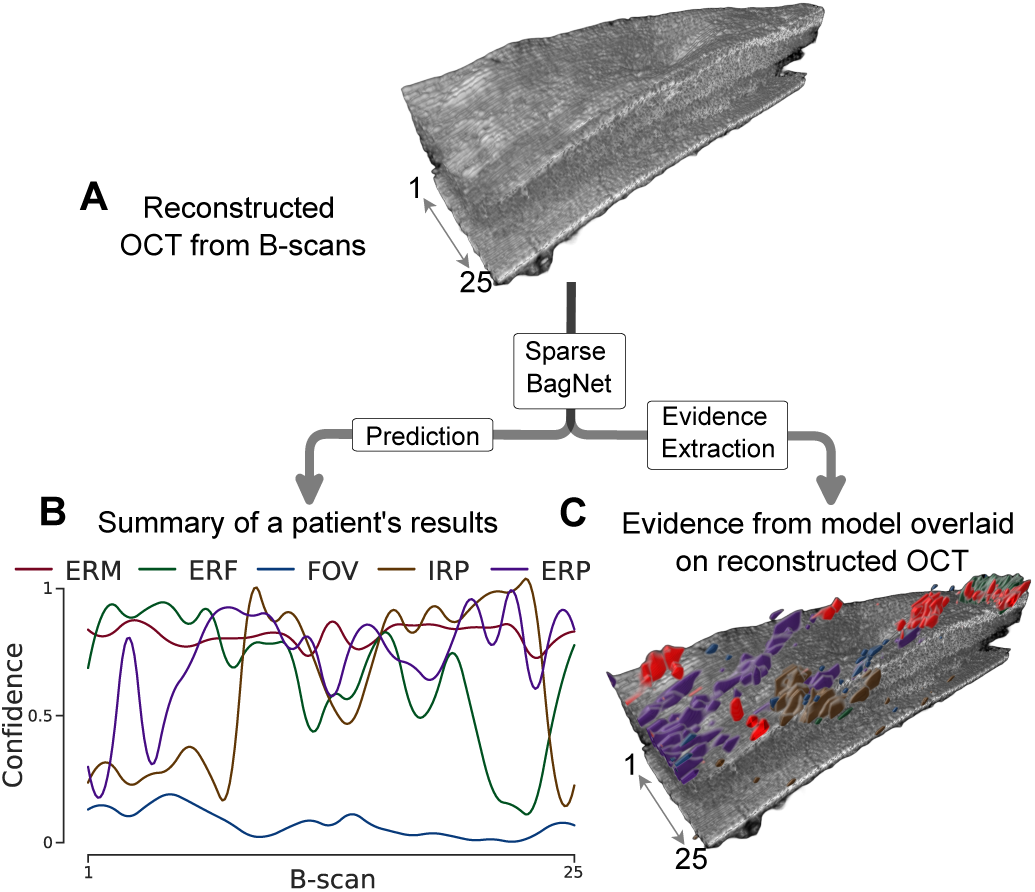
Showing an overview of the study. We input 2D B-scan OCT images to the Sparse BagNet model to obtain predictions, along with identifying disease evidence. (**A**) A 3D reconstruction from 25 B-scan images. (**B**) The results of a patient’s predictions in a summary. (**C**) Evidence generated by the model overlaid on the reconstructed OCT. The colours correspond to the pathologies.

While the ResNet–50 model demonstrated higher accuracy, it required post-hoc techniques like Grad-CAM for interpretability. These tend to produce less precise and coarse visual explanations. In comparison, the dense BagNet and the sparse BagNet models achieved comparable performance with the added benefit of inherent interpretations. The dense BagNet provided fine-grained evidence maps, but the sparse BagNet refined focus on disease-relevant regions, focusing predominantly on the top of the retina. This is essential for ensuring that the model’s output aligns with clinical expectations. Also, our sensitivity analysis indicates that the sparse BagNet model effectively highlights regions relevant for ERM predictions.

Validation on an external dataset (OCTDL) highlighted the model’s generalisation capabilities. Despite the differences in imaging devices and data distribution, the sparse BagNet model maintained high accuracy and provided interpretations, further underscoring its potential clinical utility. Moreover, the multitask adaptation of the sparse BagNet further extended its clinical relevance by reducing the need for separate models for each pathology.

The results of the user study further validate the clinical utility of the sparse BagNet model. The high level of agreement among clinicians in assessing model-generated heatmaps and patches supports the applicability of sparse BagNet in clinical decision-making. This suggests that the model’s interpretability aligns well with human expert assessment and reinforces its potential adoption in a real-world setting.

In conclusion, the sparse BagNet presents an interpretable, accurate, and clinically relevant solution for ERM detection in OCT images. It achieves this while balancing between accuracy and interpretability. Its success in both single- and multitask settings, coupled with its ability to generalise well across different datasets, highlights its potential as a valuable tool in the detection of retinal diseases.

## Code and data availability

The code used for this paper is available here https://github.com/berenslab/ERM. The datasets generated and analysed during the current study are not publicly available due to privacy reasons but may be available from the corresponding author on reasonable request.

## Declaration of Interests

The authors declare no competing interests.

## Data Availability

All data produced in the present study are available upon reasonable request to the authors.

## Acknowledgments

This project was supported by the Hertie Foundation and by the Deutsche Forschungsgemeinschaft under Germany’s Excellence Strategy with the Excellence Cluster 2064 “Machine Learning – New Perspectives for Science”, project number 390727645. PB is a member of the Else Kröner Medical Scientist Kolleg “ClinbrAIn: Artificial Intelligence for Clinical Brain Research”. We would like to express our sincere gratitude to the following clinicians for their valuable contributions to the user study: Caroline Gassel, Daniel Wenzel, Asli Giriftinoglu, Buse Gürcan and Thoko Zungu.

## Supplementary Figures

**Figure S1:**
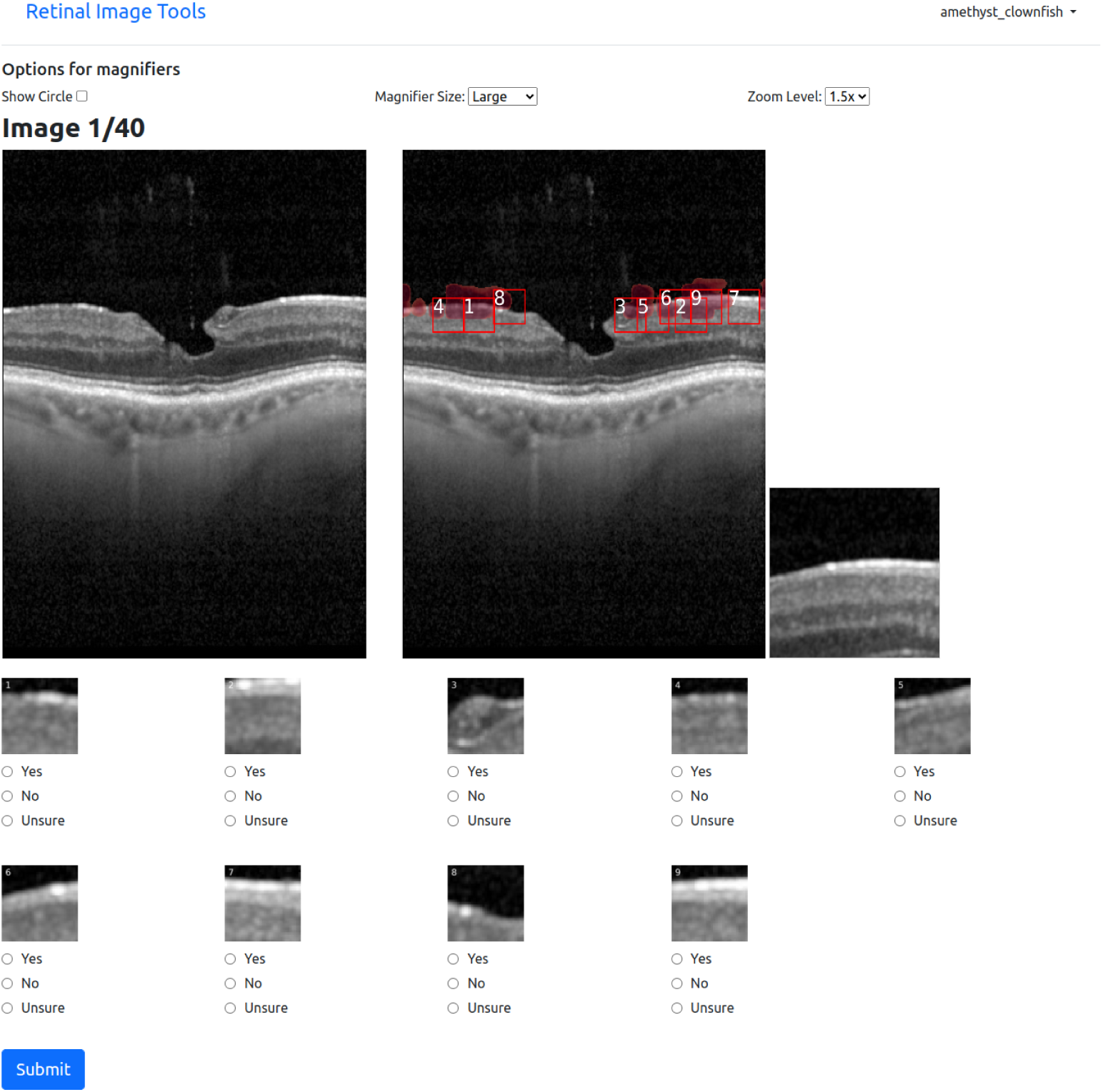
User interface for ERM evaluation. The left panel displays an OCT image and the right panel shows the same image with overlaid evidence in the form of heatmaps and patches generated by the model as potential regions of interest. Below, the patches are extracted and presented separately with participants asked to evaluate them whether they contain evidence of ERM by selecting “Yes,” “No,” or “Unsure.”

**Figure S2:**
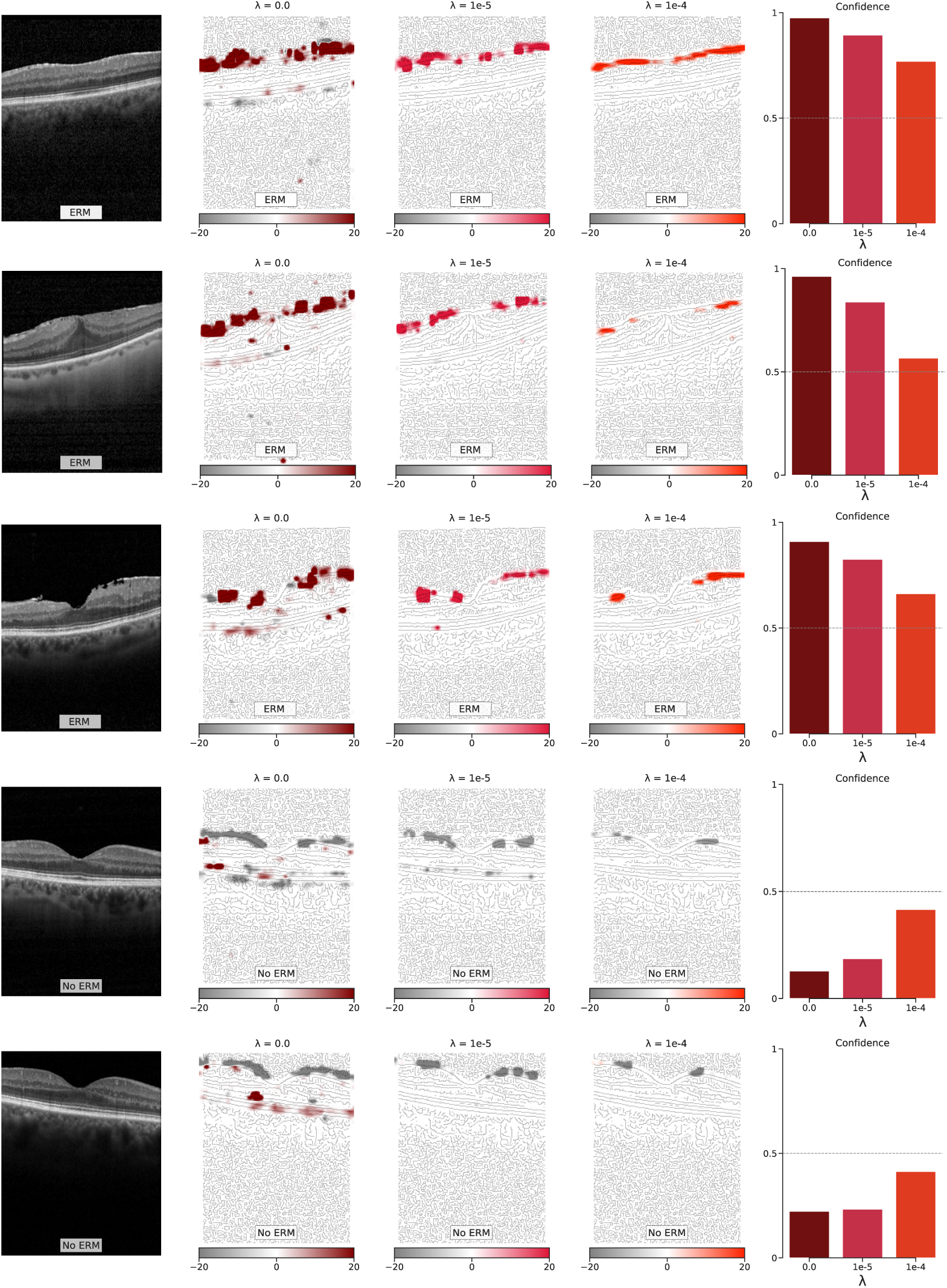
Evidence maps of varying sparsity constraints with their corresponding confidence. Additional examples demonstrating the sparse BagNet model’s detection of ERM in OCT B-scans, shown under varying sparsity constraints and their corresponding heatmaps highlighting disease-relevant regions.

**Figure S3:**
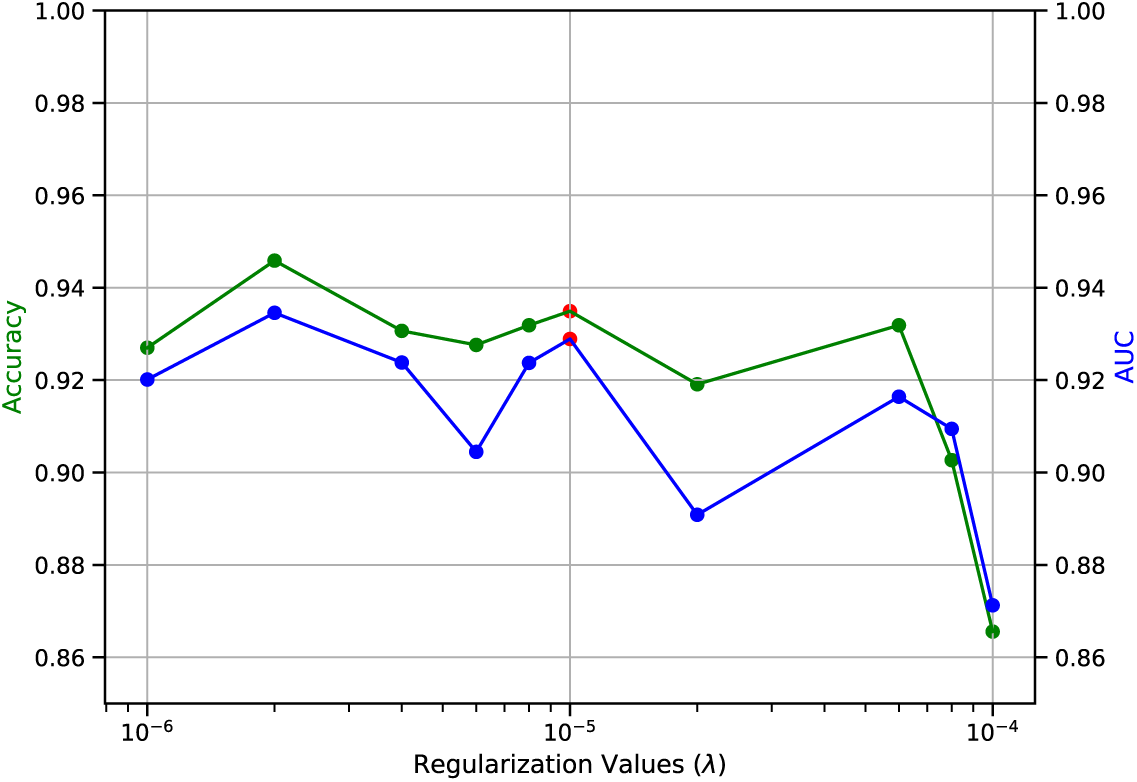
The effects of regularisation (*λ*) value on sparse BagNet performance. The red marker (*λ* = 1*×*10^−5^) represents the selected regularisation value, which is a trade-off for performance and sparsity.

**Figure S4:**
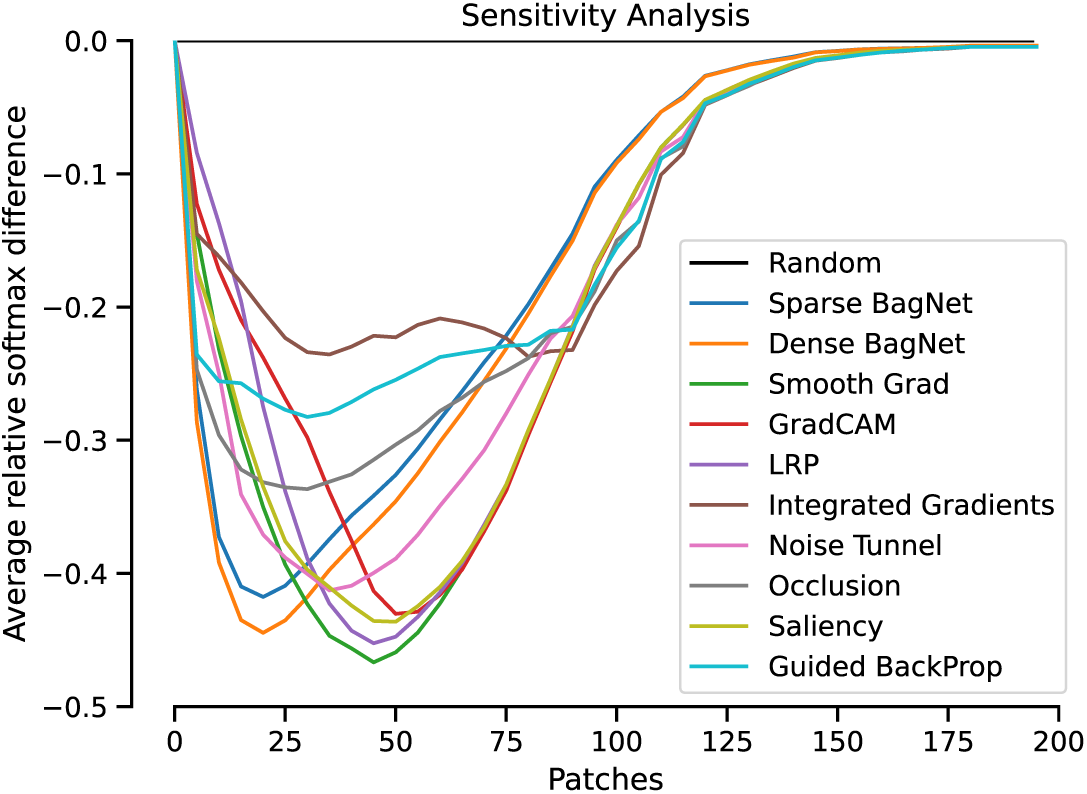
A sensitivity analysis of multiple attribution maps. The curves represent the average softmax differences from a random baseline. Patches are progressively removed to highlight the effectiveness of identifying important regions in detecting ERM in OCT images by the sparse BagNet model.

**Figure S5:**
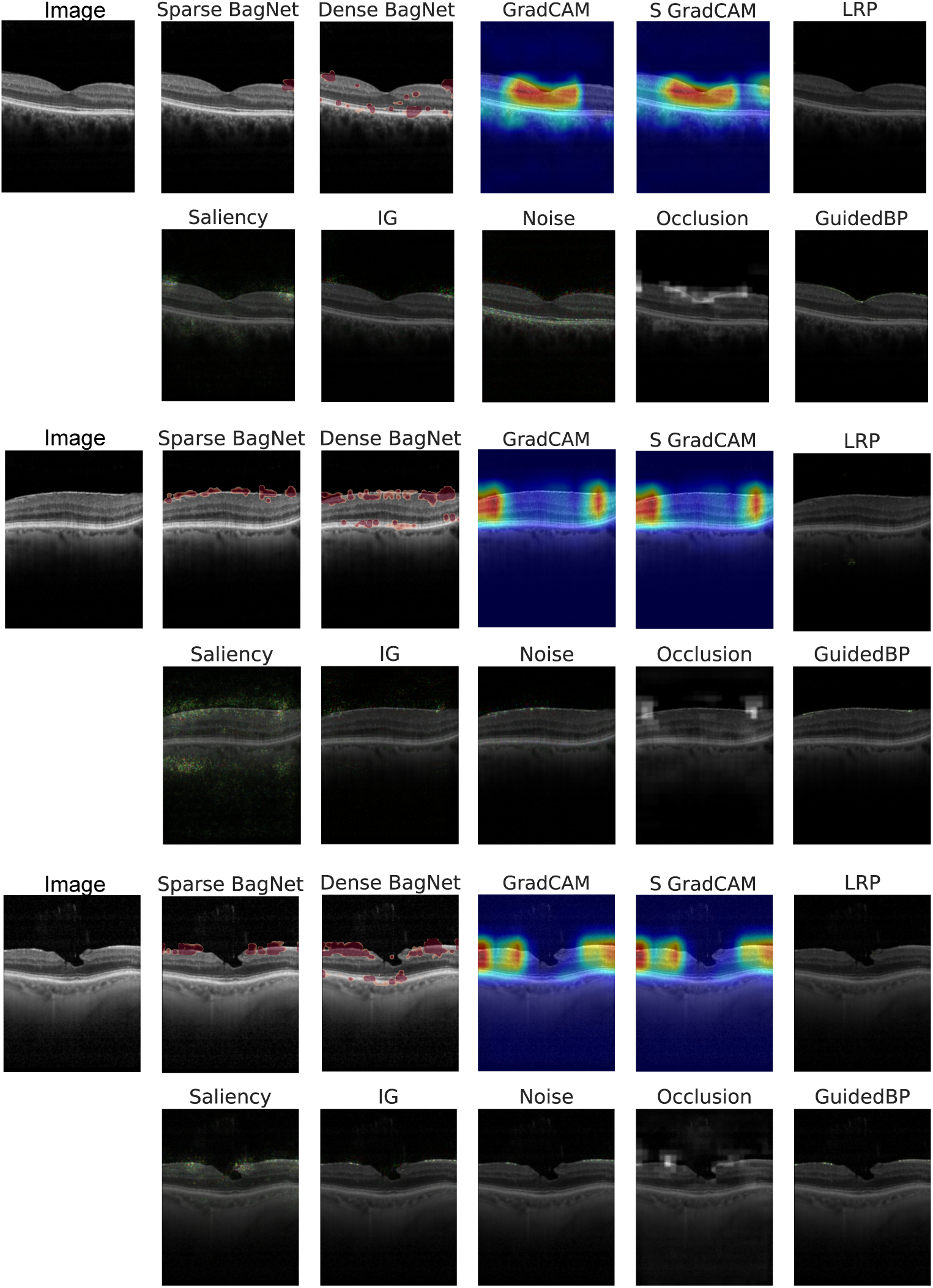
Comparing different attribution maps for ERM detection. The BagNet variants produces fine heatmaps, particularly highlighting disease-relevant regions of the OCT B-scan image – especially for the sparse BagNet. In contrast, the GradCAM variants yields coarse heatmaps. Although, the remaining attribution methods also created evidence at disease-relevant regions, these indications are smaller and less visually distinct. Moreover, they are all post-hoc interpretable techniques.

**Figure S6:**
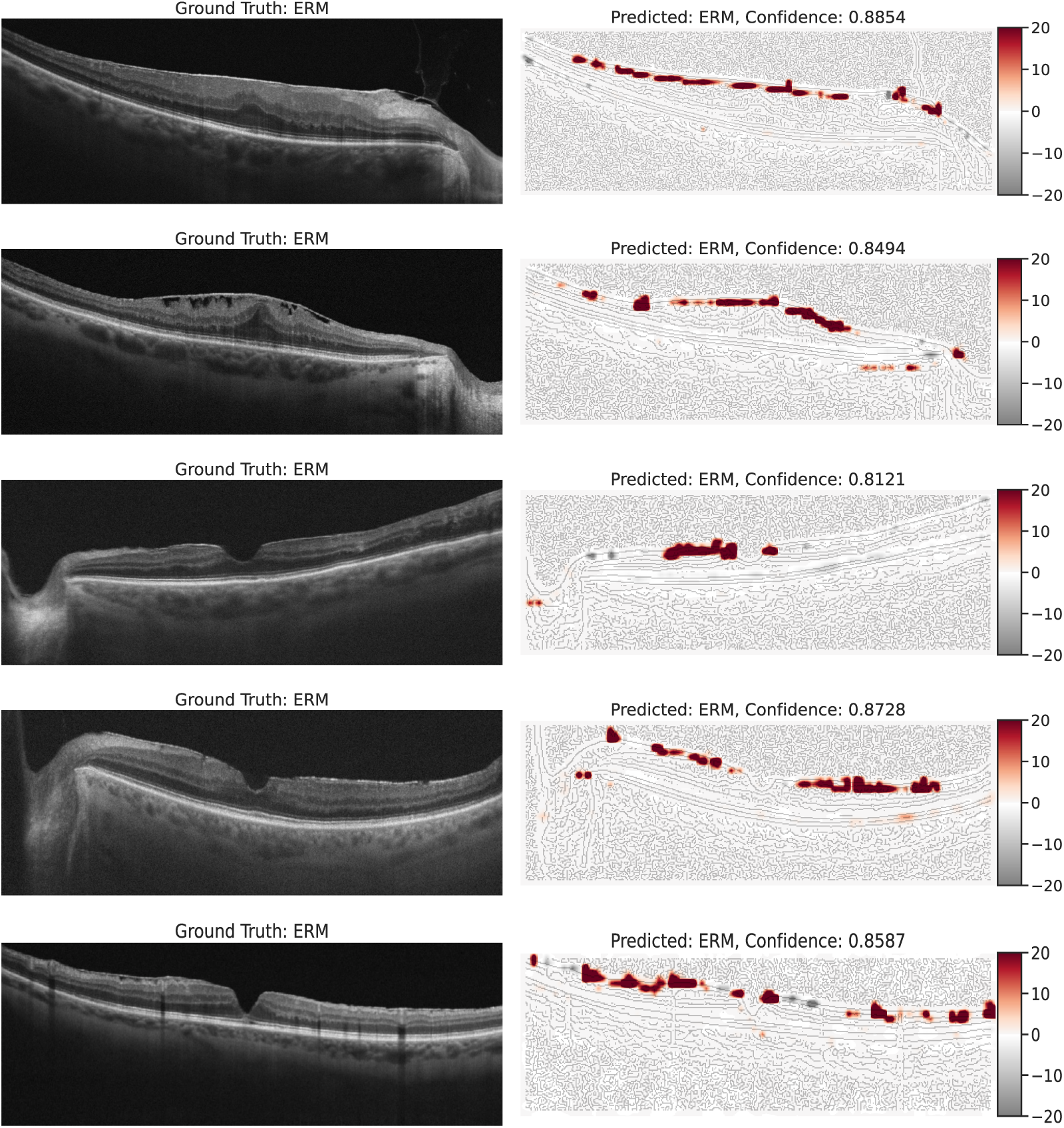
The model’s evaluation on an external dataset. Additional examples of the sparse BagNet model’s evaluation on an external dataset, with heatmaps highlighting disease-relevant areas of the OCT B-scan indicative of ERM.

**Figure S7:**
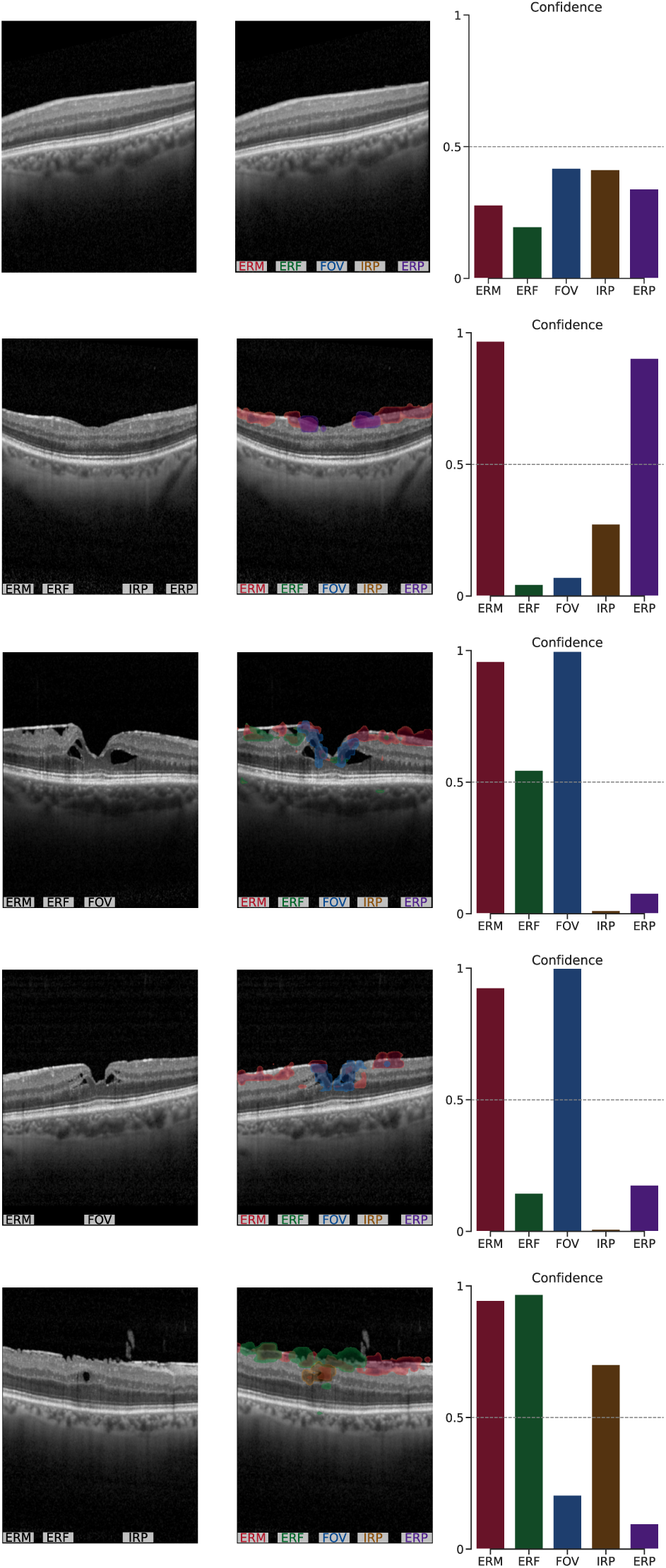
Showing several examples of visualisation of multitask sparse BagNet. The model both correctly predicts the diseases and highlights suspected regions of the OCT image.

**Figure S8:**
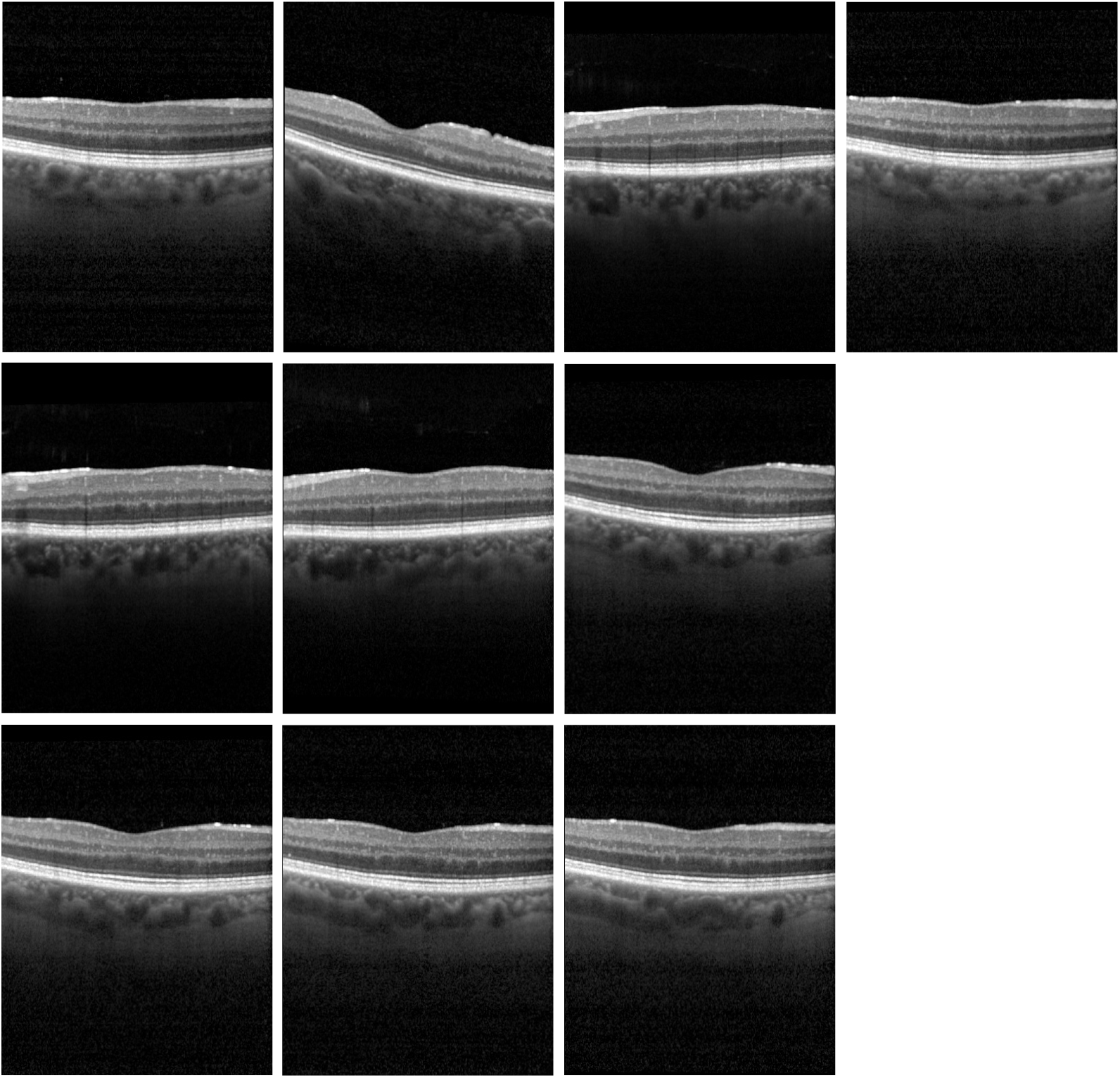
Showing the 10 incorrectly predicted OCT B-scans images used in the user study. Clinicians agreed with the models’ evidence of ERM, suggesting subtle features may have been overlooked during labelling.

## Notes

### Competing Interest Statement

The authors have declared no competing interest.

### Funding Statement

This project was supported by the Hertie Foundation and by the Deutsche Forschungsgemeinschaft under Germany's Excellence Strategy with the Excellence Cluster 2064 "Machine Learning -- New Perspectives for Science", project number 390727645.

### Author Declarations

Our dataset consisted of 624 OCT volume scans from 624 eyes of 461 patients presenting to the Department of Ophthalmology at the University of Tuebingen. The study was conducted according to the guidelines and standards of the Declaration of Helsinki, and was approved by the Ethics Committee of the University of Tuebingen, Germany.

## References

[1] Aung, K.Z., Makeyeva, G., Adams, M.K., Chong, E.W.T., Busija, L., Giles, G.G., English, D.R., Hopper, J., Baird, P.N., Guymer, R.H., et al., 2013. The prevalence and risk factors of epiretinal membranes: the melbourne collaborative cohort study. Retina 33, 1026–1034.

[2] Ayhan, M.S., Faber, H., Kühlewein, L., Inhoffen, W., Aliyeva, G., Ziemssen, F., Berens, P., 2023. Multitask learning for activity detection in neovascular age-related macular degeneration. Translational Vision Science & Technology 12, 12–12.

[3] Ayhan, M.S., Neubauer, J., Uzel, M.M., Gelisken, F., Berens, P., 2024. Interpretable detection of epiretinal membrane from optical coherence tomography with deep neural networks. Scientific Reports 14, 8484.

[4] Bae, J.H., Song, S.J., Lee, M.Y., 2019. Five-year incidence and risk factors for idiopathic epiretinal membranes. Retina 39, 753–760.

[5] Binder, A., Montavon, G., Lapuschkin, S., Müller, K.R., Samek, W., 2016. Layer-wise relevance propagation for neural networks with local renormalization layers, in: Artificial Neural Networks and Machine Learning–ICANN 2016: 25th International Conference on Artificial Neural Networks, Barcelona, Spain, September 6-9, 2016, Proceedings, Part II 25, Springer. pp. 63–71.

[6] Bradski, G., 2000. The opencv library. Dr. Dobb’s Journal: Software Tools for the Professional Programmer 25, 120–123.

[7] Brendel, W., Bethge, M., 2019. Approximating CNNs with bag-of-local-features models works surprisingly well on imagenet. arXiv preprint arXiv:1904.00760.

[8] Bu, S.C., Kuijer, R., Li, X.R., Hooymans, J.M., Los, L.I., 2014. Idiopathic epiretinal membrane. Retina 34, 2317–2335.

[9] Caruana, R., 1997. Multitask learning. Machine learning 28, 41–75.

[10] Chehaibou, I., Pettenkofer, M., Govetto, A., Rabina, G., Sadda, S.R., Hubschman, J.P., 2020. Identification of epiretinal proliferation in various retinal diseases and vitreoretinal interface disorders. International journal of retina and vitreous 6, 1–9.

[11] De Fauw, J., Ledsam, J.R., Romera-Paredes, B., Nikolov, S., Tomasev, N., Blackwell, S., Askham, H., Glorot, X., O’Donoghue, B., Visentin, D., et al., 2018. Clinically applicable deep learning for diagnosis and referral in retinal disease. Nature medicine 24, 1342–1350.

[12] Djoumessi, K.D., Ilanchezian, I., Kühlewein, L., Faber, H., Baumgartner, C., Bah, B., Berens, P., Koch, L., 2023. Sparse activations for interpretable disease grading.

[13] Djoumessi Donteu, K.R., Huang, Z., Kuehlewein, L., Rickmann, A., Simon, N., Koch, L.M., Berens, P., 2024. An inherently interpretable AI model improves screening speed and accuracy for early diabetic retinopathy. medRxiv, 2024–06.

[14] Fung, A.T., Galvin, J., Tran, T., 2021. Epiretinal membrane: a review. Clinical & Experimental Ophthalmology 49, 289–308.

[15] Ganaie, M.A., Hu, M., Malik, A.K., Tanveer, M., Suganthan, P.N., 2022. Ensemble deep learning: A review. Engineering Applications of Artificial Intelligence 115, 105151.

[16] Gende, M., de Moura, J., Novo, J., Ortega, M., 2022. End-to-end multi-task learning approaches for the joint epiretinal membrane segmentation and screening in OCT images. Computerized Medical Imaging and Graphics 98, 102068.

[17] Govetto, A., Lalane III, R.A., Sarraf, D., Figueroa, M.S., Hubschman, J.P., 2017. Insights into epiretinal membranes: presence of ectopic inner foveal layers and a new optical coherence tomography staging scheme. American journal of ophthalmology 175, 99–113.

[18] Hanumunthadu, D., Keane, P.A., Balaskas, K., Dubis, A.M., Kalitzeos, A., Michaelides, M., Patel, P.J., 2021. Agreement between spectral-domain and swept-source optical coherence tomography retinal thickness measurements in macular and retinal disease. Ophthalmology and therapy 10, 913–922.

[19] He, K., Zhang, X., Ren, S., Sun, J., 2016. Deep residual learning for image recognition, in: Proceedings of the IEEE conference on computer vision and pattern recognition, pp. 770–778.

[20] Hsia, Y., Lin, Y.Y., Wang, B.S., Su, C.Y., Lai, Y.H., Hsieh, Y.T., 2023. Prediction of visual impairment in epiretinal membrane and feature analysis: A deep learning approach using optical coherence tomography. The Asia-Pacific Journal of Ophthalmology 12, 21–28.

[21] Huang, D., Swanson, E.A., Lin, C.P., Schuman, J.S., Stinson, W.G., Chang, W., Hee, M.R., Flotte, T., Gregory, K., Puliafito, C.A., et al., 1991. Optical coherence tomography. science 254, 1178–1181.

[22] Huff, D.T., Weisman, A.J., Jeraj, R., 2021. Interpretation and visualization techniques for deep learning models in medical imaging. Physics in Medicine & Biology 66, 04TR01.

[23] Iuliano, L., Fogliato, G., Gorgoni, F., Corbelli, E., Bandello, F., Codenotti, M., 2019. Idiopathic epiretinal membrane surgery: safety, efficacy and patient related outcomes. Clinical Ophthalmology, 1253–1265.

[24] Jin, K., Yan, Y., Wang, S., Yang, C., Chen, M., Liu, X., Terasaki, H., Yeo, T.H., Singh, N.G., Wang, Y., et al., 2023. iERM: An interpretable deep learning system to classify epiretinal membrane for different optical coherence tomography devices: A multi-center analysis. Journal of Clinical Medicine 12, 400.

[25] Kanukollu, V.M., Agarwal, P., 2023. Epiretinal membrane, in: StatPearls [Internet]. StatPearls Publishing.

[26] Kingma, D.P., 2014. Adam: A method for stochastic optimization. arXiv preprint arXiv:1412.6980.

[27] Kraus, M.F., Potsaid, B., Mayer, M.A., Bock, R., Baumann, B., Liu, J.J., Hornegger, J., Fujimoto, J.G., 2012. Motion correction in optical coherence tomography volumes on a per a-scan basis using orthogonal scan patterns. Biomedical optics express 3, 1182–1199.

[28] Kulyabin, M., Zhdanov, A., Nikiforova, A., Stepichev, A., Kuznetsova, A., Ronkin, M., Borisov, V., Bogachev, A., Korotkich, S., Constable, P.A., et al., 2024. OCTDL: Optical coherence tomography dataset for image-based deep learning methods. Scientific Data 11, 365.

[29] Kunavisarut, P., Supawongwattana, M., Patikulsila, D., Choovuthayakorn, J., Watanachai, N., Chaikitmongkol, V., Pathanapitoon, K., Rothova, A., 2022. Idiopathic epiretinal membranes: visual outcomes and prognostic factors. Turkish Journal of Ophthalmology 52, 109.

[30] Li, X., Xiong, H., Li, X., Wu, X., Zhang, X., Liu, J., Bian, J., Dou, D., 2022. Interpretable deep learning: Interpretation, interpretability, trustworthiness, and beyond. Knowledge and Information Systems 64, 3197–3234.

[31] Lo, Y.C., Lin, K.H., Bair, H., Sheu, W.H.H., Chang, C.S., Shen, Y.C., Hung, C.L., 2020. Epiretinal membrane detection at the ophthalmologist level using deep learning of optical coherence tomography. Scientific reports 10, 8424.

[32] Matoba, R., Morizane, Y., 2024. Epiretinal membrane: an overview and update. Japanese Journal of Ophthalmology, 1–11.

[33] Mohammed, A., Kora, R., 2023. A comprehensive review on ensemble deep learning: Opportunities and challenges. Journal of King Saud University-Computer and Information Sciences 35, 757–774.

[34] Ng, C.H., Cheung, N., Wang, J.J., Islam, A.F., Kawasaki, R., Meuer, S.M., Cotch, M.F., Klein, B.E., Klein, R., Wong, T.Y., 2011. Prevalence and risk factors for epiretinal membranes in a multi-ethnic united states population. Ophthalmology 118, 694–699.

[35] Pang, C.E., Spaide, R.F., Freund, K.B., 2014. Epiretinal proliferation seen in association with lamellar macular holes: a distinct clinical entity. Retina 34, 1513–1523.

[36] Parra-Mora, E., Cazaũas-Gordon, A., Proeņca, R., da Silva Cruz, L.A., 2021. Epiretinal membrane detection in optical coherence tomography retinal images using deep learning. IEEE Access 9, 99201–99219.

[37] Paszke, A., Gross, S., Massa, F., Lerer, A., Bradbury, J., Chanan, G., Killeen, T., Lin, Z., Gimelshein, N., Antiga, L., et al., 2019. PyTorch: An imperative style, high-performance deep learning library. Advances in neural information processing systems 32.

[38] Pedregosa, F., Varoquaux, G., Gramfort, A., Michel, V., Thirion, B., Grisel, O., Blondel, M., Prettenhofer, P., Weiss, R., Dubourg, V., Vanderplas, J., Passos, A., Cournapeau, D., Brucher, M., Perrot, M., Duchesnay, E., 2011. Scikitlearn: Machine learning in Python. Journal of Machine Learning Research 12, 2825–2830.

[39] Randolph, J.J., 2005. Free-marginal multirater kappa (multirater *κ*_free_): An alternative to fleiss’ fixed-marginal multirater kappa. Online submission.

[40] Rasheed, K., Qayyum, A., Ghaly, M., Al-Fuqaha, A., Razi, A., Qadir, J., 2022. Explainable, trustworthy, and ethical machine learning for healthcare: A survey. Computers in Biology and Medicine 149, 106043.

[41] Rudin, C., 2019. Stop explaining black box machine learning models for high stakes decisions and use interpretable models instead. Nature machine intelligence 1, 206–215.

[42] Salahuddin, Z., Woodruff, H.C., Chatterjee, A., Lambin, P., 2022. Transparency of deep neural networks for medical image analysis: A review of interpretability methods. Computers in biology and medicine 140, 105111.

[43] Selvaraju, R.R., Cogswell, M., Das, A., Vedantam, R., Parikh, D., Batra, D., 2017. Grad-CAM: Visual explanations from deep networks via gradient-based localization, in: Proceedings of the IEEE international conference on computer vision, pp. 618–626.

[44] da Silva, R.A., Roda, V.M.d.P., Matsuda, M., Siqueira, P.V., Lustoza-Costa, G.J., Wu, D.C., Hamassaki, D.E., 2022. Cellular components of the idiopathic epiretinal membrane. Graefe’s Archive for Clinical and Experimental Ophthalmology 260, 1435–1444.

[45] Singh, A., Sengupta, S., Lakshminarayanan, V., 2020. Explainable deep learning models in medical image analysis. Journal of imaging 6, 52.

[46] Singh, G., Yow, K.C., 2021. These do not look like those: An interpretable deep learning model for image recognition. IEEE Access 9, 41482–41493.

[47] Smilkov, D., Thorat, N., Kim, B., Viégas, F., Wattenberg, M., 2017. SmoothGrad: removing noise by adding noise. arXiv preprint arXiv:1706.03825.

[48] Springenberg, J.T., Dosovitskiy, A., Brox, T., Riedmiller, M., 2014. Striving for simplicity: The all convolutional net. arXiv preprint arXiv:1412.6806.

[49] Stevenson, W., Prospero Ponce, C.M., Agarwal, D.R., Gelman, R., Christoforidis, J.B., 2016. Epiretinal membrane: optical coherence tomography-based diagnosis and classification. Clinical ophthalmology, 527–534.

[50] Sundararajan, M., Taly, A., Yan, Q., 2017. Axiomatic attribution for deep networks, in: International conference on machine learning, PMLR. pp. 3319–3328.

[51] Tang, Y., Gao, X., Wang, W., Dan, Y., Zhou, L., Su, S., Wu, J., Lv, H., He, Y., 2023. Automated detection of epiretinal membranes in OCT images using deep learning. Ophthalmic Research 66, 238–246.

[52] Teng, Q., Liu, Z., Song, Y., Han, K., Lu, Y., 2022. A survey on the interpretability of deep learning in medical diagnosis. Multimedia Systems 28, 2335–2355.

[53] Tjoa, E., Guan, C., 2020. A survey on explainable artificial intelligence (xai): Toward medical xai. IEEE transactions on neural networks and learning systems 32, 4793–4813.

[54] Yan, Y., Huang, X., Jiang, X., Gao, Z., Liu, X., Jin, K., Ye, J., 2024. Clinical evaluation of deep learning systems for assisting in the diagnosis of the epiretinal membrane grade in general ophthalmologists. Eye 38, 730–736.

[55] Zeiler, M.D., Fergus, R., 2014. Visualizing and understanding convolutional networks, in: Computer Vision–ECCV 2014: 13th European Conference, Zurich, Switzerland, September 6-12, 2014, Proceedings, Part I 13, Springer. pp. 818–833.

[56] Zhang, H., Cisse, M., Dauphin, Y.N., Lopez-Paz, D., 2017. mixup: Beyond empirical risk minimization. arXiv preprint arXiv:1710.09412.

